# A hybrid algorithm for dental artifact detection in large computed tomography datasets

**DOI:** 10.1101/2020.08.06.20169516

**Authors:** Colin Arrowsmith, Reza Reiazi, Mattea L. Welch, Michal Kazmierski, Tirth Patel, Aria Rezaie, Tony Tadic, Scott Bratman, Benjamin Haibe-Kains

## Abstract

Computed tomography (CT) is one of the most common medical imaging modalities and the main technology used in radiomics research, the computational voxel-level analysis of medical images. Analysis of CT images is vulnerable to the effects of dental artifacts (DA) caused by metal implants or fillings. Running automated analysis pipelines with uncurated datasets can reduce performance and hamper future reproducibility on new datasets. This work introduces a new tool to detect the location and magnitude of DAs in CT images based on a combination of deep learning and conventional image processing algorithms. We show the utility of this new DA detector through an analysis of the correlations between radiomic features and the location of DAs in 2,319 CT axial volumes. We were able to predict the correct DA magnitude (no, weak or strong artifacts) yielding a Matthews correlation coefficient of 0.73 (p-value=0.0002), achieving the same level of agreement as human labellers. The algorithm was able to identify the location of the DAs in the CT volumes with performance on par with human labellers. Finally, our analysis of radiomic features showed that only when strong DAs were present, the proximity of the tumour to the mouth was highly correlated with specific radiomic features. Our results suggest that removing these features, or removing CT slices containing the DAs, could reduce these unwanted correlations.

## INTRODUCTION

Computed tomography (CT) images are a commonly-used medical imaging modality. Recent advances in machine learning and deep learning have led to the development of advanced image processing techniques for medical imaging applications, including CT scans [1]. CT-derived quantitative features (also referred to as radiomic features) have shown promising results in personalized medicine [2], and when combined with machine learning, have potential utility in diagnostic and prognostic applications. Unfortunately, these radiomic features may be highly sensitive to high-density materials such as metal prosthesis or dental fillings [3]; the latter commonly causes dental artifacts, which pose a problem for imaging of head and neck patients. The metal in dental fillings has a much larger atomic number than soft tissues, resulting in a significantly higher attenuation for x-ray beams passing through the metal. As a result, these dental artifacts (DA) present as bright and dark streaks on the reconstruction images. These artifacts not only obscure large portions of the image’s reconstructed pixels, but studies have also shown that dental artifacts alter features computed by radiomics computational platforms in CT images [3,4]. They also affect target volume delineation [5], and radiation therapy dose calculation accuracy [6]. There is a need to account for artifacts during image data processing.

Several studies have tried to address this data processing challenge by removing slices affected by DAs [3] or by using metal artifact reduction (MAR) algorithms [7]. Recently, a convolutional neural network (CNN) [8] and hand-crafted radiomic feature-based model [9] have been developed to detect the presence of DAs in CT volumes. However, to the best of our knowledge, no studies differentiated between DAs of different magnitudes or quantified how the location of these artifacts could affect quantitative imaging features used to train radiomic models. Furthermore, previous DA detection studies have classified hand-drawn regions of interest (ROI) as DA positive or DA negative [9] but have not examined the correlation between radiomic features in a given ROI and its distance from the DA source. These methods, even if effective at screening datasets for artifacts, could cause vast amounts of data to be unnecessarily marked as unclean, even if the artifacts do not homogeneously affect radiomic features in the patient’s image volume.

In this study, we introduce Detection of Artifacts for Automated Radiomics Technologies (DAART), a new algorithm to detect DAs on a per-slice basis in CT image datasets. Conventional image processing methods based on histogram-based thresholding and the CT sinogram are combined with a CNN network in order to create an automated three-class DA classifier and DA location detector for large radiomic datasets. Finally, we found a small set of radiomic features, all using the same filter, to be correlated with the physical distance between the DA and the gross tumour volume (GTV).

## METHODS

The design of our study is represented in Figure 1.

**Figure 1.**
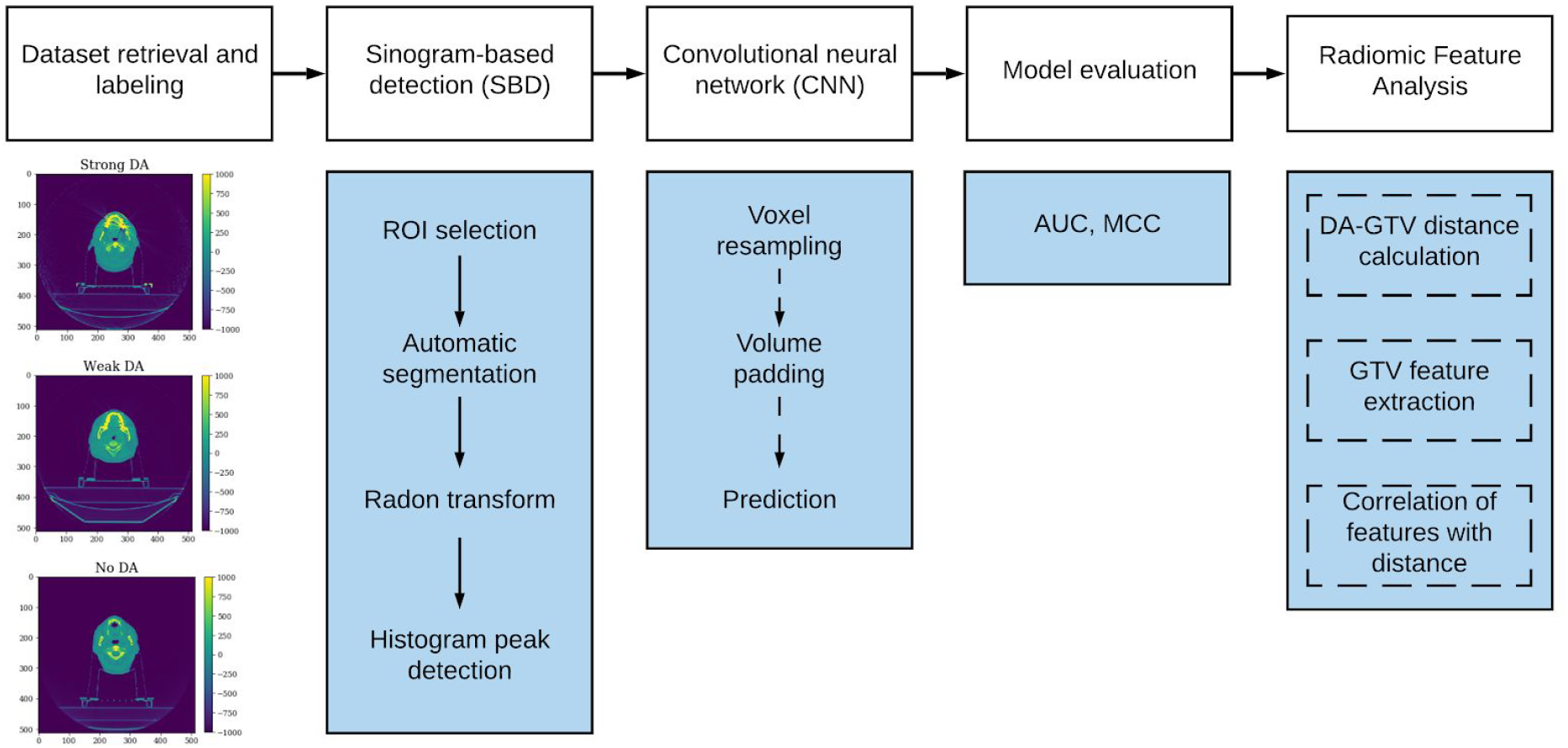
The study design includes five main steps: (1) retrieval of head and neck CT imaging volume dataset and labelling of DA; (2) initial classification of DA using a sinogram-based detection (SBD) method; (3) secondary classification of SBD-classified dental artifacts using a previously trained CNN; (4) model evaluation; and (5) exploration of the effect of DA magnitude and its distance from the GTV on radiomic features.

### Dataset retrieval and labelling

The dataset used for this study consists of 3,211 head and neck cancer CT image volumes collected from 2005-07-26 to 2017-08-17 at the University Health Network (UHN) in Toronto, Canada (REB approval #17-5871). This dataset is referred to as *RADCURE*. Each patient’s CT volume contained a median of 181 slices, with each slice consisting of 512 × 512 pixels. The median slice thickness varied between 2.0 and 3.0 mm for different patients and had a median of 2.0 mm (see Supplementary Table 1 for a comprehensive list of imaging settings).

We developed Artifact Labelling Tool for Artifact Reduction (ALTAR), an open-source web-application enabling the review of large sets of images and the annotation of the magnitude and location of the dental artifacts. The application clipped all images to be between −1000 and 1000 Hounsfield Units (HU), thereby facilitating viewing of the grayscale images in a dynamic range conducive to DA identification. Manual annotations classified each of the patients as either having “strong” artifacts, “weak” artifacts, or no visible artifacts for the entire image stack. A patient labelled as “strong” had to have at least one slice containing artifact streaks which obscured significant portions of the patient’s body and which were easily visible outside the profile of the body. A patient labelled as “weak” had to have at least one slice with easily identifiable metal artifact streaks, but which did not fully obscure sections of the image and which were not plainly visible outside the patient’s body. Finally, a patient labelled as “none” had no identifiable dental artifacts. The z-index of the axial slice containing the strongest artifacts was also annotated for each patient in the strong or weak DA class. For patients with no DA, an axial slice index in the mouth was labelled. Other metal artifact streaks caused by catheters, metal implants, etc. were not taken into consideration for our study. These non-dental artifacts may be less common, as one study found that of 1,300 patients, 131 (10%) had artifacts below the head and neck region [10]. The images in our study were labelled by four different researchers, each labelling a subset of the data. The process was supervised by a researcher with 10 years of experience (details about the annotation accuracy and the analysis are reported in Supplementary Information).

### Automated Classification

Classification is achieved using a two-step process. All head and neck CT image volumes are initially classified on the basis of whether or not they contain a dental artifact using a sinogram based detection (SBD) method. The original head and neck CT image volumes are then fed through a pre-trained CNN for secondary binary dental artifact classification. By observing the combined binary classifications of the two methods, a three-class dental artifact label is determined.

#### Sinogram Based Detection (SBD)

We used a thresholding and sinogram-based approach to create an initial binary DA classifier and DA location detector for patient CT volumes. All images were cropped by taking the first 350 pixels in the x-axis to remove the majority of the imaging table and fixture accessories which occupied the last 100-200 pixels of the x-axis. The image intensity range was then clipped to between −1000 and 1000 HU and divided by 1000 HU, normalizing all images to a range of (−1, 1). Next, the head was segmented in each two-dimensional slice of the patient’s CT volume. This was done using the Otsu threshold [11] which minimizes the intra-class variance, *σ_m_*, defined below:

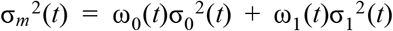

where ω_0_*(t)* and ω_1_(t) are the probabilities of the two intensity classes separated by the threshold *t* and 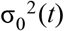 and 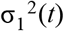 are the variances of each class. The Otsu threshold was computed using the Python scikit-image package (version 0.16.2) [12]. The resulting mask of the patient’s body and the head is extended outward by applying a Gaussian filter to the mask, followed by thresholding the filtered mask at a value of 0.01. This threshold value was chosen based on the width of the Gaussian filter kernel, to extend the edges of the body. Finally, each normalized image slice was multiplied by the mask, creating a set of two-dimensional images containing only the original pixels in the background. These images were then thresholded at a value of 0.04.

Each two-dimensional slice was then transformed into a sinogram by calculating 180 parallel projections using the Radon transform in Python’s scikit-image. Finally, a sinogram is made by adding up projections from different angles. The mean pixel intensity in a central region of each slice’s sinogram is then computed (Figure 2).

**Figure 2:**
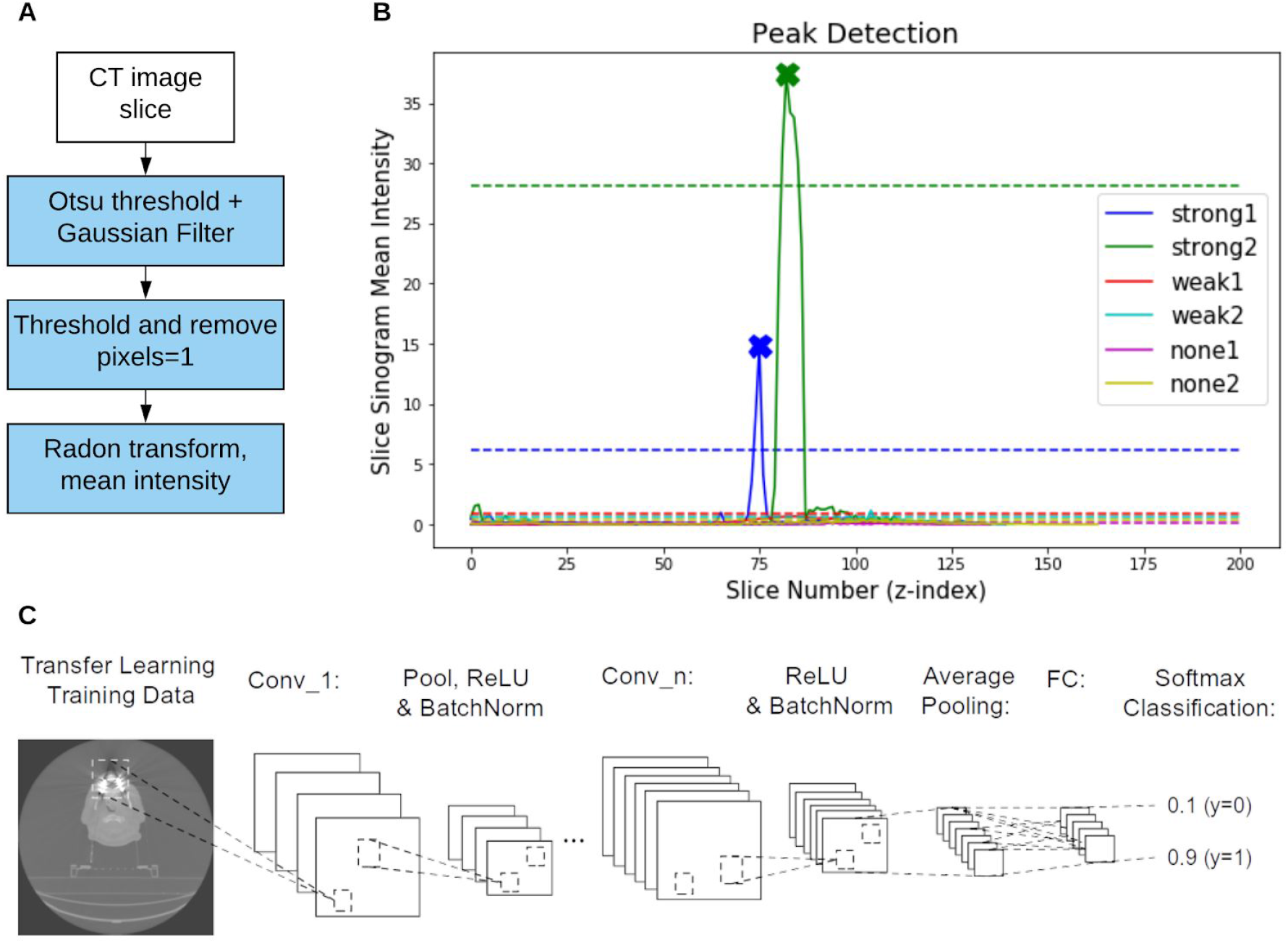
An illustration of the two binary DA classifiers used in this study. (A) Two steps in the sinogram-based detection (SBD). First, one slice from a CT volume is thresholded and blurred, before being thresholded again to remove pixels in the body of the patient. The remaining pixels are thresholded again, revealing the streaks outside the patient’s body. The image is then transformed to the sinogram domain and the mean sinogram pixel intensity is computed. (B) An example of the ‘mean sinogram intensity’ for each slice in six CT volumes (each image represented with a different colour). A peak detection algorithm is applied to this plot for a given patient to detect slices likely to contain DAs. We annotate the detected slices with Xs to show that the algorithm detected one peak from each of the green and blue curves (both images labelled as ‘strong DA’). The dashed lines represent the peak detection threshold for each patient. (C) The CNN architecture used in the study. The network consisted of 5 convolutional layers (conv_1 to conv_5) creating a total of 64 filters.

The thresholding, filtering, and sinogram steps resulted in a list of mean sinogram intensities for each axial CT slice in a given patient. We used the peak detection algorithm from the Python scipy package (version 1.4.1) to detect slices with intensities much higher than the mean for that patient. A peak is detected if it has a higher value than its neighbours and if it has an intensity more than four standard deviations above the mean (Figure 2B). The algorithm classifies the patient as DA positive if any peaks in the z-axis sinogram intensity curve are found. If no peaks are found, the patient is classified as DA negative. This method additionally outputs a prediction of the z-index of the DA when a patient CT volume is classified as DA positive.

#### Convolutional Neural network Detection

A five-layer convolutional neural network (CNN) [8] was used as a secondary classification step in our method. This CNN is a binary DA classifier that takes 3D patient CT volumes as inputs (Figure 2C). The volumes are preprocessed to contain isotropic voxels of 1×1×1 mm. The image volumes are then padded and resampled to matrix size of 256×256×256, retaining the image aspect ratio.

#### Thresholding-based DA Location Detection

A thresholding-based algorithm was developed to detect the axial slice containing a DA in a patient CT image volume. The algorithm assumes that an artifact is present in the patient. This method is intended to be used after a binary DA classifier has already classified a given patient as DA positive. This method simply attempts to iteratively decrease the thresholding of an image until the bright pixels which cause artifacts are detected.

The thresholding-based algorithm works by first clipping the intensity values between the maximum intensity in one patient’s CT image volume and 200 HU above that maximum. The standard deviation of each axial slice is then computed and peak detection is performed on the standard deviations of each axial slice using the scipy find_peaks function in a similar manner to the SBD peak detection step. A peak, defined as a slice with a standard deviation higher than its adjacent slices, is detected if it has a value *σ_i_* > μ*_V_* + 1.5*σ_V_*, where μ*_V_* is the mean of the slice standard deviations for that patient volume and *σ_V_* is the standard deviation of the slice standard deviations for that patient. If any peaks are detected, the algorithm simply returns the indices of those peaks for that patient. Otherwise, the lower bound of the clipping range is decreased by 50 HU and the process is repeated for the patient until at least one peak is found.

#### Combined SBD-CNN Three-Class DA Classifier

Classification of dental artifacts was performed using our two-step algorithm combining a sinogram-based detection (SBD) method with a convolutional neural network (CNN). Each image volume was classified by both the SBD and the CNN. A decision tree was used to classify images into no, weak and strong artifact categories (Figure 3). In other words, if an image volume is classified as DA positive by the SBD and DA positive by the CNN, it is classified as “strong”. If an image is classified as DA negative by the SDB and DA positive by the CNN it is classified as “weak”.

**Figure 3:**
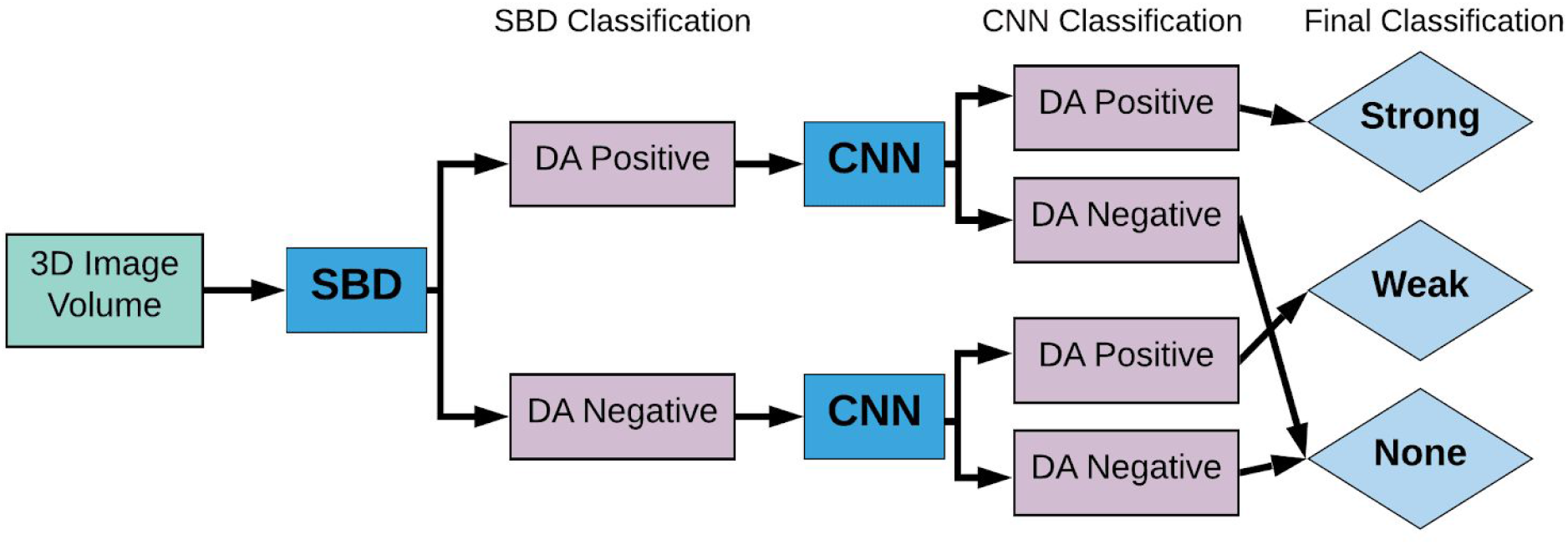
Flowchart of the SBD-CNN hybrid algorithm for dental artifact detection. Images were annotated manually and then first binned using SBD (Sinogram based detection) based on the average intensity of the corresponding sinogram. Subsequently, the original images were classified using the CNN model. Images that were labelled as artifact positive by both the SBD and CNN were categorized as having strong dental artifacts. Images labelled as artifact negative by both methods were labelled as having no artifacts. This way our hybrid model is capable of labelling images based on the strength of artifact presence.

### Radiomic Feature Analysis

The relationship between quantitative imaging features and the existence and location of dental artifacts was studied. Radiomic features were extracted using the default settings of the open-source Python package, PyRadiomics (version 2.1.2) [13,14]. Features were extracted from the GTV of each patient in the dataset. We rescaled all features such that each feature had zero mean and unit variance across all patients. This reduces the inter-feature variation due to different units, allowing us to compare only the distributions of data between features. A GTV mask was created for each patient based on contours generated by radiation oncologists. The mask was then applied to the original image, setting all pixels outside of the GTV to zero. Amongst the full set of 3,211 image volumes, 2,490 contained a GTV. The GTVs from these volumes were extracted and used to compute 1,547 radiomic features using the default parameters of the PyRadiomic package.

The axial (z-axis) slice index of the dental artifact was manually labelled for each patient as described above. The z-index of the DA was defined to be the slice with the strongest visible artifact streaks, or the most central slice in cases where no slice was obviously brightest. In cases where no DA was present, the z-index of the mouth was labelled. All images and GTV masks were resampled to 1×1×1 mm voxel spacing and the physical z-distance between the DA centre and GTV centre was calculated. The location of the GTV pixel nearest to the DA slice was also extracted for further analysis.

The difference in the distribution of radiomic feature values between strong DA and no DA images with respect to DA-GTV distances, was evaluated. The image volumes were grouped into 40mm DA-GTV distance bins and a Wilcoxon rank-sum test between image features from volumes with strong DAs and image volumes with no DAs was performed. We used the Bonferroni correction for adjusting the nominal p-values of each correlation for multiple testing. A Bonferroni-corrected p-value < 0.05 was considered significant. The relationship between DA-GTV z-distance and the radiomic features was investigated using all 2,490 patient volumes containing GTVs. For each feature, the partial Spearman correlation (adjusted for tumor volume) between the feature values and DA-GTV distances was computed independently for each DA magnitude (1,039 strong, 751 weak, 877 none). This was repeated for both measures of DA-GTV distance (GTV centre of mass and GTV pixel nearest to the DA).

### Performance Assessment

We primarily used two metrics to assess the accuracy of our prediction models.: the Matthews correlation coefficient (MCC) and the Area Under the Receiver Operating Characteristic Curve (AUC). The MCC is equivalent to the Chi-square coefficient for binary labels and accounts for the potential class imbalance. We found the MCC to be a useful metric for DA detection, as an effective dataset cleaning tool should have a high accuracy *and* a low false-negative rate (the rate at which a classifier fails to detect DAs when they are present). The MCC can also be generalized to multiclass cases, allowing us to compare the performance of our binary and three-class classifiers.

The class-weighted AUC was used to assess the accuracy of the CNN binary classifier based on the prediction scores produced by the network. The roc_auc_score function from Scikit-learn (version 0.22.1) was used with the ‘weighted’ option [15]. This computes the metrics for each class, weighted by the number of true instances for each label. For all AUC and MCC values we also estimated a p-value from 5,000 iterations of a randomized permutation test.

### Research Reproducibility

The application we created to manually annotate images, Artifact Labelling Tool for Artifact Reduction (ALTAR) can be downloaded from our GitHub repository (https://github.com/bhklab/ALTAR). The code for the SBD, CNN, and thresholding location detection is open source (Creative Commons Non-Commercial) and freely available from on our DAART GitHub repository (https://github.com/bhklab/DA-Detection). To ensure full reproducibility of our study we created a Code Ocean capsule to allow users to easily run and reuse our analysis pipeline (https://codeocean.com/capsule/2097894/tree).

## RESULTS

### Dataset Labelling

After reviewing 3,211 image sets using our ALTAR web-application, we identified 2,180 volumes containing artifacts (1,289 strong and 891 weak) and 1,031 volumes with no dental artifacts. The manual labelling was consistent in the set of 420 images that were labelled by two researchers, yielding three-class Matthews correlation coefficient (MCC) of 0.73 (p-value=0.0002), and 0.91 (p-value=0.0002) for binary classes (strong/weak vs none; Supplementary Table 2). The annotators labelled the same slice as containing the “strongest DA,” or the patient’s mouth in DA negative cases, in 46% of patients (Figure 4A). The two annotators labelled the strongest DA slice to within 5 slices of each other 82% of the time.

**Figure 4.**
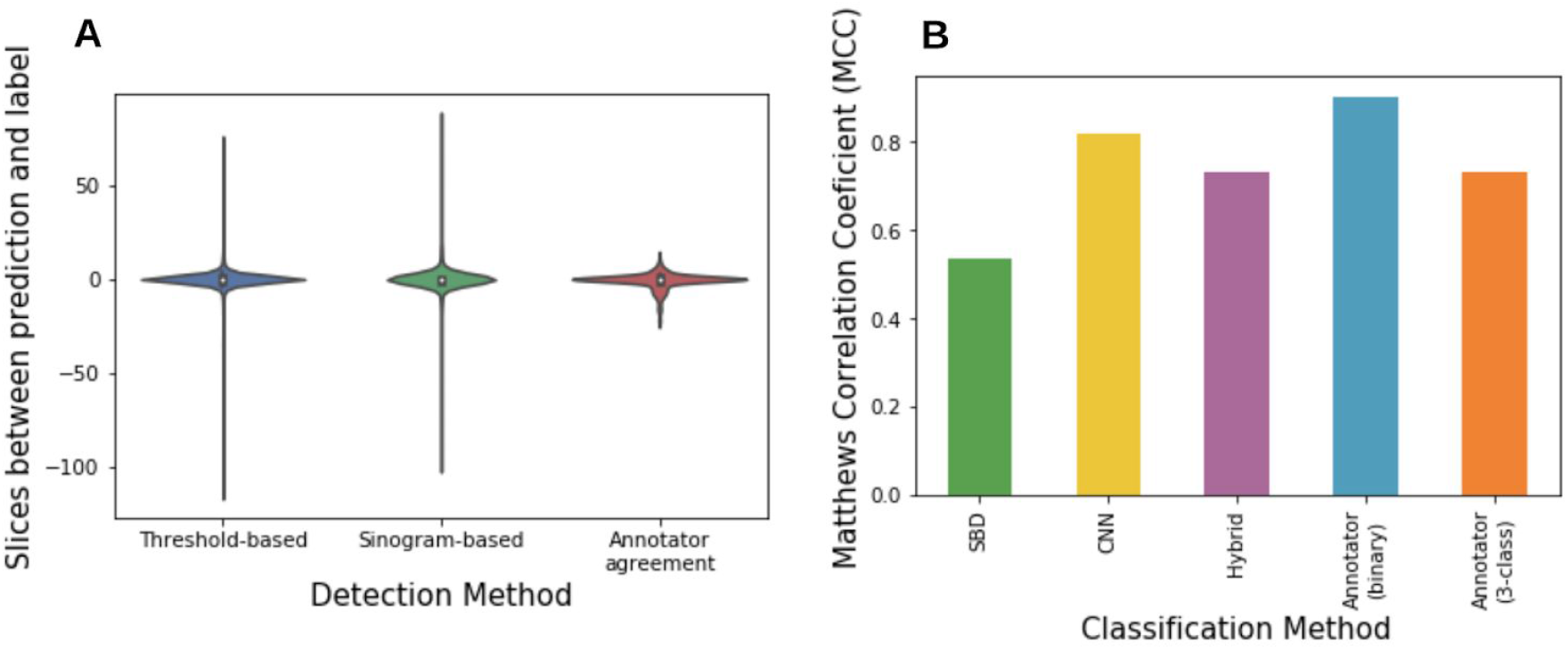
Performance of DA classification. (A) Distributions of how close the predicted slice index is to the labelled index for the threshold-based and sinogram based-detection methods (e.g. 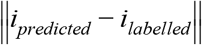). The difference in slice label between two human annotators for a set of 482 CT volumes is also shown. (B) Performance (MCC) of the DA magnitude classification techniques used in this study. The p-value of the MCC for all classifiers was < 0.001). The sinogram-based detection (SBD) and convolutional neural network (CNN) are both binary classifiers. The SBD was tested on 3,211 CT image volumes and the CNN binary classifier was tested on a subset of 2,319 image volumes. The SBD-CNN hybrid algorithm is a three-class classifier and the three-class MCC is therefore displayed here.

### Classification

#### Sinogram Based Detection (SBD)

The sinogram-based method had an accuracy (correct detection rate) of 90.5% for the strong DA image volumes and an accuracy of 24.9% for the weak DA image volumes. Combining the three DA magnitude classes to binary labels (DA positive, DA negative) the sinogram method had an overall true positive rate of 63.6%, a false positive rate of 6.7%, and a false negative rate of 36.4% (AUC=0.78, p-value=0.00002 and MCC=0.531, p-value=0.0002; Figure 4B). In images that were classified as DA positive by the algorithm, 44.5% of DA location predictions exactly matched the labelled slice index. In 88.8% of cases, the predicted location was within 5 slices of the labelled location, 92.3% were within 10 slices of the label, and 94.6% were within 15 slices of the label (Figure 4A). The median slice thickness varied between 2.0 and 3.0 mm for different patients and had a median of 2.0 mm.

#### Convolutional Neural Network Detection

The published CNN [8] was tested on 2,319 CT image volumes (945 strong, 606 weak, and 768 without artifacts). These 2,319 images are a subset of the full 3,211 image volume set that are independent from the training of the published CNN. When used on its own to make binary classifications (DA positive or DA negative) of entire patient CT volumes, the CNN yielded an MCC of 0.82 (p-value=0.0002; Figure 4B) and an AUC of 0.97 (p-value=0.0002; Supplementary Figure 3), in line with the performance of the CNN found in the original study (an AUC of 0.92 ± 0.03) [8].

#### Thresholding-based DA Location Detection

The thresholding-based DA location detection algorithm was tested on 2,087 images with artifacts (1,231 strong, 856 weak). The algorithm identified the exact slice which was labelled in 36% of cases. In 92% of cases, the algorithm identified a slice within 5 slices above or below the label (Figure 4A).The combined SBD and CNN DA three-class classifier yielded an MCC of 0.73 (p-value=0.0002) on 2,319 images (945 strong, 606 weak, and 768 without artifacts). This was identical to the three-class agreement between human annotators (MCC=0.73, p-value=0.0002; Figure 4B). This hybrid algorithm was able to make use of two different binary DA detection algorithms which independently performed worse than human labelling. Together, the two methods complement each other and are able to stratify images into three distinct DA magnitude classes.

### Analysis of Radiomic Features and Dental Artifacts

#### Analysis of the difference between patients with strong and no DAs

To test for differences between features from DA positive and DA negative image volumes, performed a Wilcoxon rank sum test between features from each group. We found that 442 features were significantly different between the DA and no DA groups (Wilcoxon rank sum test corrected p-value < 0.05), while 55 features varied significantly between strong DA and no DA patients when the artifact was 40 mm to 80 mm from the GTV. No features were significantly different between strong DA and no DA for patients with a DA more than 80 mm from the GTV.

In order to validate the effect of removing “bad” images on radiomic features, we removed all images where the centre slice of the DA overlapped with any pixel in the GTV. We performed the same Wilcoxon rank sum test between radiomic features from strong-DA and no-DA images from this smaller group of 1006 patients (529 strong, 477 no DA). We found that only 123 features were significantly different between the groups (p < 0.05). In order to verify if this reduction was simply due to inflated p-values as a result of a smaller sample size, we repeated the analysis 1000 times, each time taking a random sample of 1006 patients from the full dataset. The mean of this repeated test was 188 significant features and the value of 123 features was in the bottom 4.1% of repeated test results.

#### Analysis of the correlation between DA-GTV distance and Features

To assess the correlation between the radiomic features and distance of the GTV from the DA slice, we computed the partial Spearman correlation between DA-GTV distance and radiomic feature value, controlling for tumor volume (Figure 5). 36 features were correlated with distance only in images with strong DAs (e.g. those same features were not correlated with distance when computed from weak and no-DA images). All but two of these 36 features were found to use the “lbp-3D-k” filter. Nine of these 36 features were also found to be significantly different between strong-DA and no-DA images in the Wilcoxon rank sum test.

**Figure 5:**
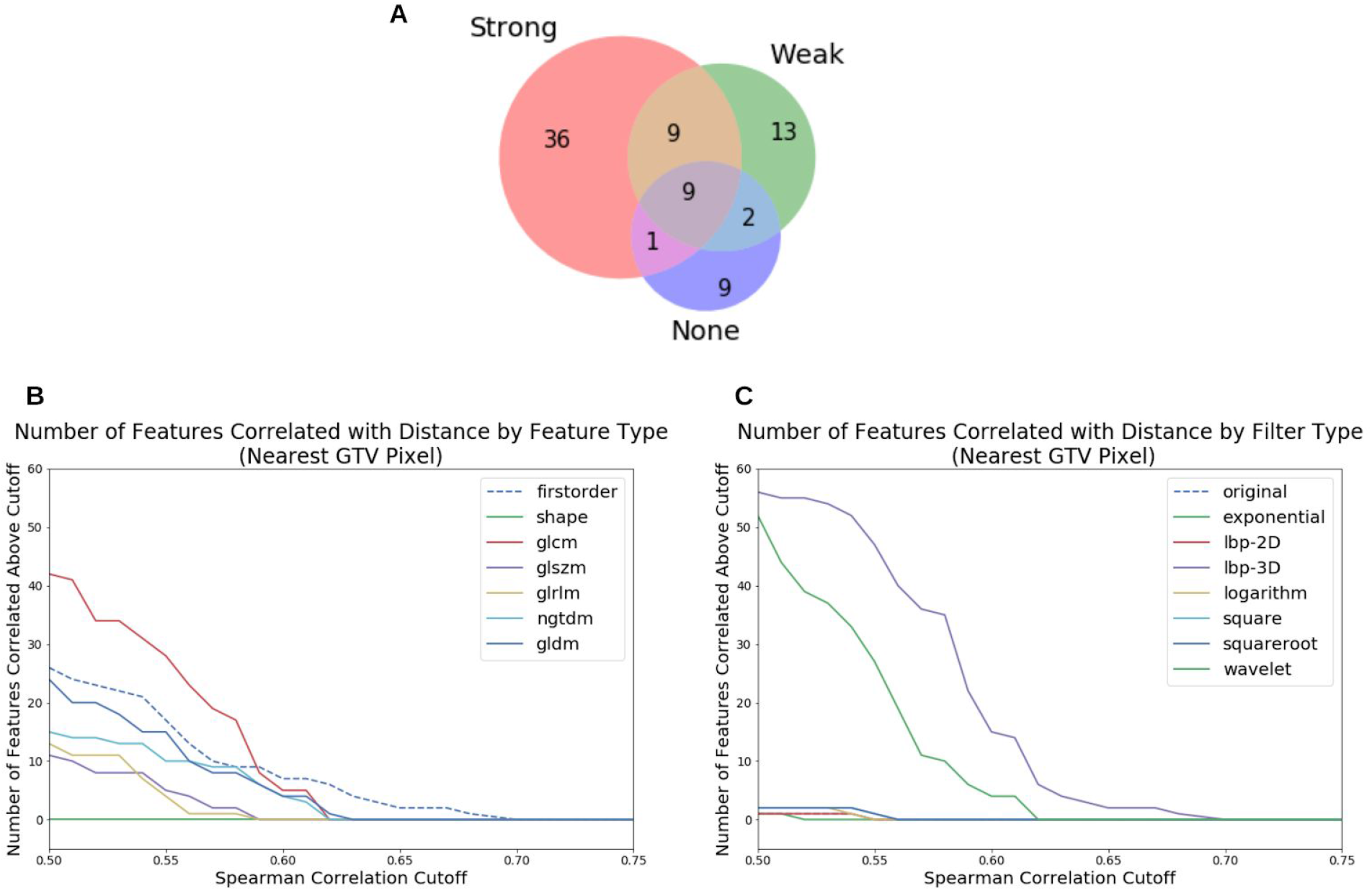
Correlation between GTV-DA distance and feature values, based on the partial correlation using Spearman correlation. (A) Venn diagram showing the number of features with |r| > 0.55 calculated from patients from each DA class. This diagram only includes significant correlations (p < 0.05). For instance, 36 features had |r| > 0.55 and were found in patients with strong DAs (pink region), but those features had |r| < 0.55 when calculated from weak or no-DA images). Nine features had |r| > 0.55 when calculated for all three DA groups (grey region). (B) The number of features with DA-GTV distance correlation above a given cutoff, grouped by feature type. (C) These correlations grouped by filter type. (B) and (C) only include significant features (p < 0.05).

## DISCUSSION

The main goal of this study consists of creating an automated pipeline for DA classification in large imaging datasets and investigating potential spurious correlations due to metal artifacts. Using a large dataset of 3,211 head and neck cancer CT image volumes we built a DA location and magnitude classifier. Using a subset of 2,490 patients, we extracted 1,547 radiomic features and investigated statistical differences in these features based on their DA locations and magnitude.

To automate DA classification, we propose a DA magnitude and location classifier to help reduce confounding correlations relating to DAs. Rather than using whole-CT volume DA classifiers to remove samples or patients from training sets, we suggest using a DA location detection algorithm to only remove images where the DA is close to the CT region of interest. The binary DA CNN classifier from Welch *et al*. was found to perform just as well on this larger dataset (AUC = 0.97, p-value=0.0002) as it had in the original study (AUC = 0.92 ± 0.03) and outperformed our sinogram-based detection method. However, we were able to leverage the SBD’s ability to discriminate between strong and weak artifacts (90% true positive rate for strong, 25% for weak DAs) to create a three-class DA classifier. The three-class classifier also performed as well as two human annotators, with both the three-class MCC of the annotators and the algorithm being 0.73 (p-value = 0.0002).

In addition to developing a novel multi-class dental artifact detection method, we developed an algorithm to detect the slice containing the artifact. We found that both the sinogram-based and the thresholding-based location detection methods agreed with a human-annotator as well as two humans would agree with one another. This was determined by comparing 482 images in our dataset that were annotated twice by different human observers (see supplementary). The algorithms predicted the DA to be within 5 slices of the human label in 80-90% of cases, while two humans agreed on the label to within 5 slices in 82% of cases. Due to the simplicity of the thresholding-based algorithm, it could be used as an efficient add-on to any DA detection algorithm, or used to annotate the locations of DAs in datasets where the DA status of images is known, but not their location. This could be useful for removing data corrupted by DAs from a dataset without having to exclude the entire CT volume of DA positive patients.

Analysis of radiomic features between strong DA and no DA patients revealed that approximately a third of features varied significantly with dental artifact status when the DA was less than 40 mm from the GTV. We found that the number of features associated with dental artifact status decreased significantly as the distance between the GTV and DA increased. This suggests that the location plays an important role in the effect of DAs on radiomic features. To further investigate this distance dependence, we examined the correlation between DA-GTV distance and the radiomic feature values. We found that, among the correlated features (Spearman correlation >0.55), only 0.5% of features (9 out of 1,547 radiomic features) were found in DA negative volumes only. However, a larger set of 36 correlated features were specifically found in the strong DA volumes. Interestingly, 34 features in this set are exclusively composed of radiomic features which used the “LocalBinaryPattern3D-kurtosis” (‘lbp-3D-k’) filter, suggesting that their lack of robustness in the presence of DA makes them unsuitable for radiomics modeling. lbp-3D-k filter computes the kurtosis (a measure of the tailedness of a distribution) [16] from the local binary pattern, a rotationally invariant measure of texture in three dimensions [17]. We hypothesize that DAs are altering the width of the distribution of specific texture metrics in three dimensions. These strong correlations between DA-GTV distance and specific radiomic features highlights the need for robust data curation pipelines for DAs in radiomic studies.

We were then able to further motivate the use of DA location detection in dataset cleaning. By removing images with their GTV in the same slice as the DA, the number of features significantly different between strong-DA and no-DA images was significantly reduced (123 DA-affected feature vs an average of 188 features by randomly selecting 1006 patients; p-value = 0.041). This highlights the need for a DA location detector in dataset cleaning. By removing only the images with a GTV overlapping with a DA, we were able to significantly improve the robustness of the features extracted from the dataset.

Interestingly, the fact that only nine of those 36 distance-correlated features were significantly different between strong-DA and no-DA images (based on the Wilcoxon rank sum test) suggests that these two analyses are detecting different types of dependency. In particular, using the Spearman correlation with GTV-DA distance may be a more strict criterion by which to select features to exclude from a radiomics study. We suggest using both analyses in order to select features robust to DAs in future radiomics studies.

Our study has several potential limitations. The analysis in this study has largely focused on the vertical location of DAs and their vertical distance from the GTV. This ignores any potential relationship between DA distance and radiomic features in the x, y plane (within a slice). We also acknowledge the inherent subjectivity of our manual labelling process, as individual researchers and clinicians may have widely varying definitions of strong, weak and no artifacts. Although our analysis of annotator agreement shows that this was not a major problem in our study, it does mean that our work could be difficult to reproduce with different data and researchers.

In conclusion, we have developed a novel dental artifact detection algorithm which when combined with a convolutional neural network, created a three-class classifier for CT images with strong, weak, and no DAs. We then created a simple thresholding-based algorithm to detect the location of DAs in DA positive CT volumes. These new tools have been made open-source to be used in future studies to assess and account for the effects of DAs on radiomic models. We stress that our findings suggest that radiomic features are affected not only by the presence of DAs, but also by their location in the images.

## Data Availability

https://github.com/bhklab/DA-Detection

https://codeocean.com/capsule/2097894/tree

## ACKNOWLEDGEMENTS

This research was supported by the Canadian Institutes for Health Research (CIHR) radiomics grant (# 426366). We would like to thank the head and neck group at the Princess Margaret Cancer Centre for their contributions to this study and their invaluable work to create the dataset used in this study. We would also like to acknowledge the contributions of the Radiomics Medicine Program at the University of Toronto and the University Health Network. Finally, we would like to thank Dean Zhu and Aleesha Masud for their help annotating the data and helping develop ALTAR.

